# The Oldenburg Hearing Health Record (OHHR)

**DOI:** 10.1101/2025.03.30.25324761

**Authors:** Sumbul Jafri, Daniel Berg, Mareike Buhl, Matthias Vormann, Samira Saak, Kirsten C. Wagener, Christiane M. Thiel, Andrea Hildebrandt, Birger Kollmeier

**Affiliations:** Department of Psychology, School of Medicine and Health Sciences, Carl von Ossietzky Universität Oldenburg, Germany; Cluster of Excellence “Hearing4All”, Carl von Ossietzky Universität Oldenburg, Germany; Department of Medical Physics and Acoustics, School of Medicine and Health Sciences, Carl von Ossietzky Universität Oldenburg, Germany; Université Paris Cité, Institut Pasteur, AP-HP, Inserm, CNRS, Fondation Pour l’Audition, Institut de l’Audition, IHU reConnect, F-75012 Paris, France; Hörzentrum Oldenburg gGmbH, Germany

## Abstract

Hearing health is shaped by both measurable auditory function and the perceived ability to navigate daily life. To fully understand its complexities, objective assessments of hearing and functional performance must be complemented by subjective reports on lived hearing experiences. The Oldenburg Hearing Health Record (OHHR) was developed to unite these measures, offering a comprehensive and open-access resource for hearing health. It contains data from 581 adults aged 18–86 years (*255 females; mean age = 67.31 years; SD = 11.93*) with varying degrees of hearing loss. Data were collected between 2013 and 2015 at the Hörzentrum Oldenburg in collaboration with Hearing4all. OHHR includes audiometric tests (Pure Tone Audiometry, Loudness Scaling, Speech in Noise tests), self-reports on hearing difficulties, lifestyle, technology use, and cognitive assessments (DemTect, Vocabulary size test). These measurements remain relevant in clinical and research settings. The dataset supports cross-disciplinary analyses linking hearing ability with cognition and quality of life, contributing to personalized hearing healthcare and advancing precision medicine.

## Background

Hearing loss is a major global health concern^1–3^. More than 1.5 billion people experience some degree of hearing loss during their lifetime, with at least 430 million requiring hearing health care. In Europe alone, approximately 190 million individuals (20% of the population) are affected to varying degrees by hearing loss or deafness^1^. Beyond its sheer prevalence, hearing loss has a profound impact on an individual’s health and quality of life. It has been associated with an elevated risk of cognitive decline, increased perception of cognitive load, and is a contributing factor to accelerated brain atrophy, particularly in regions critical for language and memory^4–7^. Neuroimaging studies show that even mild-to-moderate hearing loss increases listening effort and alters functional brain connectivity in areas supporting attention and cognitive control^8^. This increased listening effort reflects the additional cognitive resources required to process degraded auditory input and maintain speech understanding^9^.

Hearing loss also has significant psychosocial consequences, including isolation, depression, and reduced social engagement, all of which further harm well-being^10,11^. Indeed, hearing loss disrupts every aspect of daily life, from difficulties understanding speech in noisy environments to safety concerns linked to poor sound localization. The sustained effort required to listen can be exhausting, with studies showing significantly elevated fatigue levels in individuals with hearing loss^12^, often leading to social withdrawal. In many cases, the social stigma surrounding hearing aid use, and the added burden of comorbidity with tinnitus can make coping with hearing loss even more challenging. In workplace and educational settings, hearing loss can impede performance and achievement, further diminishing quality of life^13^.

All these multifaceted associations between hearing loss, psychosocial well-being, and health underscore the need for a data-driven investigation and simultaneous modelling of these multivariate associations. Such an approach requires large, shared databases that incorporate subjective reports, multiple objective auditory measures (e.g., speech-in-noise perception, loudness tolerance), assessment of cognitive performance, comorbid health conditions and psychosocial factors to improve and promote an individualized understanding of the impact of hearing loss. The promotion of comprehensive hearing health datasets will facilitate the extension of knowledge beyond the confines of traditional and small-scale audiometric assessment and provide insight into complex multivariate associations.

An overview of existing hearing data repositories was recently published by Sethukumar and colleagues (2023)^14^. Their catalogue brings together a wide range of international publications, including studies on hearing loss, ageing, neurodegenerative disorders, healthcare and disease registries, birth cohorts and population-based studies. Each study comprises one or a few limited types of hearing data, which taken together include pure-tone audiometry, speech-in-noise measurements, otoacoustic emissions, auditory brainstem response, speech discrimination, loudness recruitment, and other clinical and self-reported hearing measures. Notably, sixty percent of the studies included in the catalogue refer to objective hearing data, and in some cases, even genetic testing. The catalogue also highlights datasets with extensive audiological testing, such as the Japanese Intractable Inner Ear Databank^15^, the Swedish National Databank on Sudden Sensorineural Hearing Loss^16^, and the Childhood Development after Cochlear Implant Dataset from Johns Hopkins University, USA^17^. Some cohort studies also incorporate broader biopsychosocial data along with hearing related measures from larger populations, thus providing potential for valuable insights into hearing health.

Despite these substantial contributions to the study of hearing loss, the advancement of hearing research is hampered by the lack of comprehensive datasets that integrate extensive audiological test data with cognitive, social, lifestyle, and medical variables in a single record that is openly available for scientific discovery. Many of the studies catalogued by Sethukumar and colleagues (2023) do not have access to the participants’ lifestyle and medical history. Although the UK Biobank^18^ for example offers an extensive biomedical dataset, it lacks pure tone audiometry data and uses only the Digit Triplet Test as a screening tool for speech in noise testing, limiting its utility in evaluating functional hearing abilities. Other studies, such as the British 1970 Cohort^19^ and the AGES Reykjavik Study^20^, provide audiometric and self-reported data but do not include any speech-in-noise measurements, which are crucial for understanding real-world communication challenges. The Rotterdam Study^21^ is a longitudinal investigation of the incidence and determinants of chronic disease and disability in older adults. It combines data on hearing, cognition, physical and mental health, genetic testing, neuroimaging, and epidemiological data. However, the primary focus of this study in the field of otolaryngology is alone age-related hearing loss or presbycusis. This focus limits its applicability to other forms of hearing impairment, such as noise induced or ototoxic hearing loss and congenital conditions. The ongoing Rheinland study^22^ also has a special focus, namely on aging and neurodegenerative disorders. Both studies employ the Mini-Mental State Examination for cognitive screening, which cannot detect subtle cognitive challenges faced by individuals with hearing problems^23^. Although the standard Mini-Mental State Examination remains widely used; research indicates that the Montreal Cognitive Assessment and the Dementia Detection test are more sensitive in detecting early cognitive decline in diverse populations^24,25^. Importantly, access to many of these datasets requires formal and complex authorization, some of which are financially burdensome, and others have institutional affiliation restrictions, all of which limit their availability for widespread and accelerated research. It is worth noting, however, that some of these studies are ongoing and their policies and access options may update in the future.

To address the limitations of existing hearing health datasets and their limited accessibility, we describe and share here a comprehensive open access dataset. T h e Oldenburg Hearing Health Data Record (OHHR) was collected between 2013 and 2015 at the Hörzentrum Oldenburg gGmbH in collaboration with the Cluster of Excellence “Hearing4all” in Germany. OHHR is focused on individuals with well-defined hearing loss and comprises a hearing loss cohort of 442 out of a total of 581 participants. Having a well-defined cohort of individuals who are deaf or hard of hearing, and those who are unaffected allows researchers to isolate the effects of hearing impairment from potential confounding variables. Although other datasets, such as birth cohorts or disease-specific studies, offer valuable insights, they may not capture the same level of detail on hearing loss and associated health and well-being. The current data resource offers an extensive array of self-reported and examiner-driven measures in conjunction with pure tone audiometry, thereby providing researchers with a multifaceted perspective on the nature of hearing loss and associated factors. It also includes a precise speech-in-noise test, i.e. the Göttingen Sentence Test^26^ to characterize the individuaĺs functional hearing deficit (to be treated with a hearing device) and the Digit Triplet Test, which is widely used as a hearing screening tool for assessing speech recognition in noisy environments^27–29^. The Dementia Detection test was used to identify cognitive impairment, primarily assessing fluid cognitive abilities such as memory and attention. In addition, a vocabulary size test was administered as a proxy for crystallized intelligence and to estimate premorbid cognitive ability^30^. This distinction is particularly useful when interpreting screening outcomes, as it helps differentiate between long-standing individual differences (reflected by crystallized ability) and clinically relevant early cognitive decline (reflected by fluid ability)^31,32^. To achieve a holistic assessment, the dataset considers not only clinical evaluations but also draws from self-reported information using standardized questionnaires.

Thus, OHHR captures a broad spectrum of data, including socio-demographic characteristics, hearing health history and impact on daily life, noise exposure, hearing aid use and satisfaction, mental and physical health status, and potential comorbidities of hearing loss. Its increased granularity facilitates the exploration of the interplay between hearing loss, functional hearing deficit, cognitive decline, and overall quality of life and health.

Precision medicine in audiology undoubtedly benefits from extensive datasets such as the OHHR. This dataset has supported several significant studies within the field, demonstrating its utility in identifying individual predictors of speech-in-noise performance^33^, exploring determinants of hearing aid uptake^34^, developing Common Audiological Functional Parameters^35–37^ and advancing auditory profiling for clinical decision support systems^38–40^.

However, these publications relied on internal, unstructured versions of the dataset and did not involve all the variables or any data for public sharing. Their focus was primarily on audiological outcomes, with limited integration of the broader cognitive, psychosocial, or demographic data included in the resource. In contrast, the curated subset described in this paper has undergone extensive refinement in terms of structure, consistency, exhaustiveness and documentation. It includes comprehensive variable-level metadata, detailed test descriptions, and quality control protocols that were not publicly available before. While this release draws from the same broader data source used in prior publications; it now offers a more complete, transparent, and accessible version of the OHHR that is suitable for reuse by the wider research community.

The release of OHHR in a standardized and accessible format creates opportunities for interdisciplinary research across audiology, cognitive science, psychology, and public health. The dataset’s integration of audiological, psychosocial, cognitive, and demographic data enables a wide range of applications—from developing predictive models for personalized rehabilitation to informing strategies for hearing loss prevention and public health planning. Researchers can use OHHR to identify high-risk populations, evaluate factors influencing hearing aid adoption, and assess the broader impact of hearing loss on quality of life and daily functioning.

The inclusion of patient-reported outcomes, together with comprehensive data on hearing aid use, provides a strong foundation for advancing patient-centered care models and evaluating assistive technologies in real-world contexts. OHHR also supports the exploration of diverse factors that shape the perception and management of hearing difficulties, contributing to a more nuanced understanding of hearing loss. Its structured format and thorough documentation promote reproducibility and cross-study comparisons, making it a valuable resource for data-driven approaches in clinical and public health research. By making this curated dataset openly available, we aim to support research on hearing loss by providing resources for early detection, intervention development, and rehabilitation studies.

## Methods

### Participants

Participants aged 18 years and older were recruited from an existing database of volunteers at Hörzentrum Oldenburg gGmbH (abbreviated as HZO in the following). This non-profit organization is affiliated with the Carl von Ossietzky Universität Oldenburg and partners with the “Hearing4all” Cluster of Excellence funded by the German Research Foundation (https://hearing4all.de/en/). It works with a global network of scientific and industrial collaborators to develop innovative audiological methods and relies on the support of about 2,000 local volunteers. After screening for completeness, the final dataset prepared for public release via Zenodo (https://zenodo.org/records/14177902)^41^ includes 581 individuals, aged 18 to 86 years (*n* = 255 female; median age = 70.0 years; mean age = 67.31 years; standard deviation (*SD*) = 11.93 years). The data was anonymized in accordance with the General Data Protection Regulation (GDPR; Regulation (EU) 2016/679). Each participant was assigned a unique identifier to maintain anonymity while enabling multivariate analyses. Data protection approval for the preparation and open release of the dataset was granted by the Data protection and Information Security Management Office at the Carl von Ossietzky University of Oldenburg, Germany.

While the OHHR cohort includes a higher proportion of individuals with hearing loss than would be expected in the general population, its purpose was not to reflect population-level prevalence of hearing loss. Rather, the aim was to establish a large cohort oversampling a wide range of hearing impairments, offering a robust foundation for research focused on the deaf and hard-of-hearing individuals. This contrasts with many population-based datasets, which often include a majority of normal-hearing individuals. Additional information on the hearing loss prevalence and how the sample compares to population-level estimates is provided in the *Technical validation* section.

### Procedure

The data collection process began with an 11-page questionnaire, referred to as the “home questionnaire”, which volunteers completed at home. It was sent via postal mail to potential participants along with study information and consent forms, with instructions to complete everything using pencil and paper and return it via prepaid mail. Participants who returned the completed home questionnaire were then invited to a one-hour lab-based test session. This initial process ensured that participants understood the nature of the data they would be sharing prior to any in-person testing. The test session included a face-to-face interview to discuss the individual’s medical history, as well as several assessments. Besides a clinically standard pure tone audiogram, supra-threshold loudness perception was determined with the Adaptive Categorical Loudness Scaling^42^. In addition, two speech-recognition-in-noise tests (the Göttingen Sentence Test - GÖSA^26^ and the German Digit Triple Test - DTT^28^), and two cognitive tests (Vocabulary size test or Wortschatztest - WST^43^ and the Dementia Detection test - DemTect^25^) were performed. All measures were undertaken by trained professional staff members. Hearing assessments, including Pure Tone Audiometry, Adaptive Categorical Loudness Scaling, the GÖSA, and the German DTT, were performed without hearing aids, even for regular users. However, cognitive assessments and the questionnaires, where optimal hearing was required were conducted with hearing aids for those who typically used them^33^.

### Ethics and data anonymization

The local ethics committee at the Carl von Ossietzky Universität Oldenburg reviewed and authorized the original data collection between 2013 and 2015, ensuring compliance with the ethical standards described in the Declaration of Helsinki^44^, except for the requirement for preregistration of the study. Participants were compensated with 12€ per hour for their involvement.

To prepare the current publicly available dataset, additional ethical approvals and data protection measures were implemented. All records were anonymized to protect participant privacy. To minimize the risk of re-identification, k-anonymity (k=4) was applied, ensuring that each participant’s quasi-identifiers were indistinguishable from at least three others.

Further safeguards were introduced, including grouping categories with fewer than four cases into broader classifications. The full anonymization process was reviewed and approved by the university’s data protection officer, and documentation outlining the procedures and risk assessments are available upon request. At the time of the original data collection, approximately 40% of participants had consented to the storage of their contact information. These individuals were re-contacted and provided explicit consent for the publication of their anonymized data. For the remaining 60%, whose data had already been pseudonymized and who could not be re-contacted, a waiver of consent for data sharing was granted by the data protection officer following the completion of the anonymization process.

Data protection approval for the preparation and public release of the dataset was subsequently granted by the Data Protection and Information Security Management Office at Carl von Ossietzky Universität Oldenburg (Application Number: DSM-H4A Open Dataset/20241113-0009).

### Self-reports included in the home questionnaire (HQ)

Subjective experience of hearing loss related consequences reflects the functional impact of hearing loss on an individual’s life, an aspect that cannot be captured by audiometric measures alone. There has been much discussion in the literature about the low association between examiner-driven (“objective”) and self-reported (“subjective”) measures of hearing loss. However, it is crucial to also characterize individuals with hearing loss in terms of their subjective hearing-related experience, along with their subjective health status (chronic diseases), self-reported demographics (age, gender), personality traits (self-efficacy, self-esteem), mood (depressive symptoms) and social context (social network size)^45^. Research by Wang and colleagues (2020)^46^ show that underreporting of hearing problems is associated with age, ear problems, lifestyle, and subjective beliefs about hearing health. Notably, older adults tend to underreport their hearing difficulties^47^. A combined approach of self-reports and audiometric measures is also effective in detecting high-frequency hearing loss.

Furthermore, self-reports are particularly sensitive in contributing to the identification of mid-frequency hearing loss^48^. Understanding the factors that influence self-reported hearing problems, including cognitive abilities, education, and lifestyle, enables researchers and practitioners to interpret these reports more precisely and use them as complementary measures to audiometry for prediction and for deriving individualized treatment recommendations^49^. The HQ covered several key assessments, which are described in detail below.

#### Hearing anamnesis

The HQ collected detailed information on the individual’s hearing history, including diagnosis and duration of hearing impairment, subjective ratings of hearing problems in both quiet and noisy environments, and noise exposure. Individuals were asked about the causes of their hearing difficulties (see Figure 1), hearing aid use (past and present), duration of use, and any history of sudden sensorineural hearing loss, for example due to middle ear infections. Age-related hearing loss (ARHL) was among the most frequently reported causes. In the context of this dataset, ARHL also known as presbycusis is a bilateral, symmetrical, and progressive sensorineural hearing loss that occurs with advancing age. It is characterized by reduced hearing sensitivity, especially for high-frequency sounds, and may be accompanied by reduced speech understanding, particularly in noisy environments. Ear noise/tinnitus was also addressed. This included its occurrence, duration and the physical or mental discomfort it causes. Furthermore, to evaluate the comfort and experience of hearing aid use, the questionnaire included three items from the seven-item International Outcome Inventory for Hearing Aids^50^.

**Figure 1.**
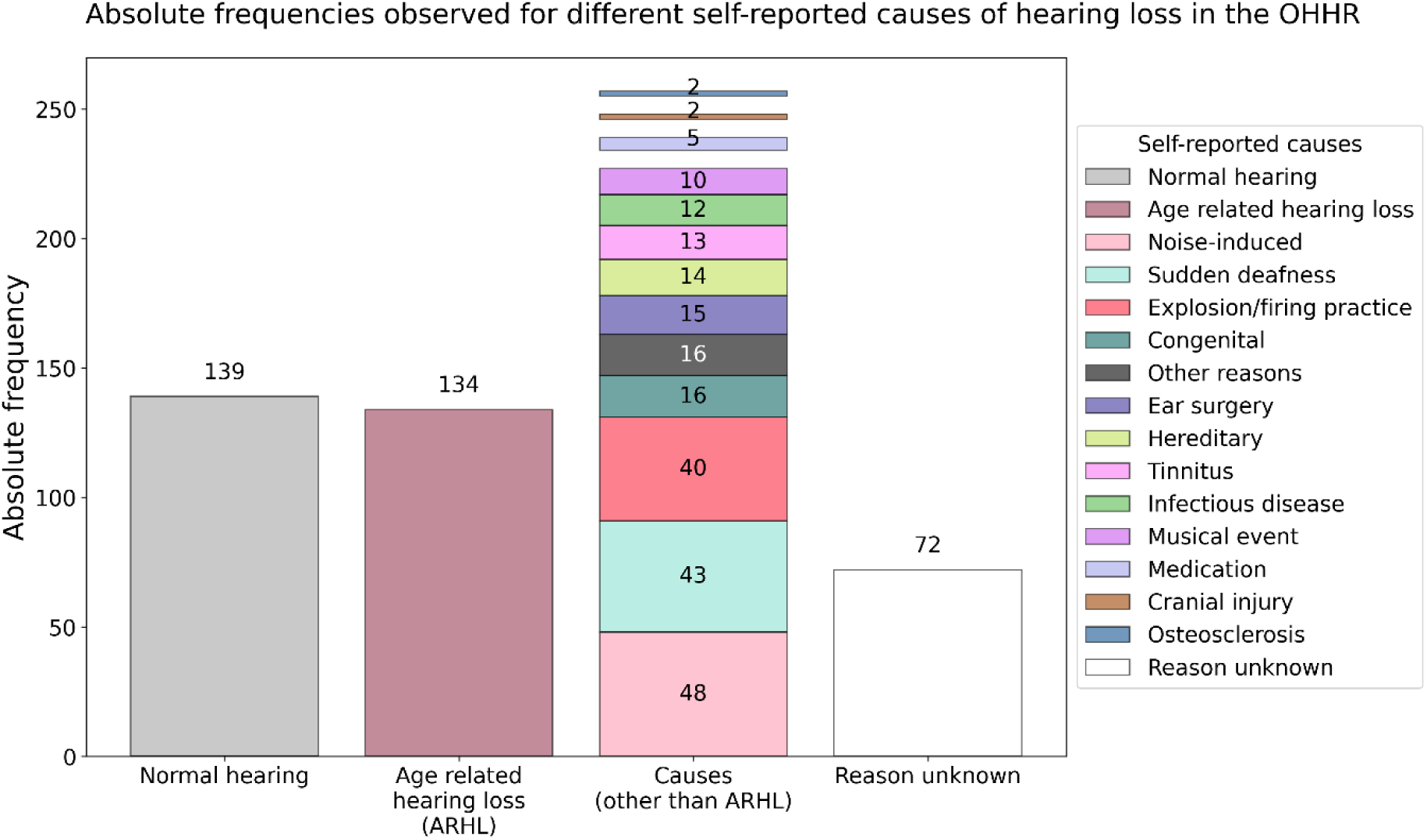
Self-reported causes of hearing difficulties. *Note*. Absolute frequencies of self-reported causes of hearing difficulties in the OHHR dataset (N = 581). A total of 370 individuals reported a perceived cause of hearing loss, while 72 reported not knowing the reason and 139 reported normal hearing. Age-related hearing loss (ARHL) or presbycusis showed the highest prevalence among the reported causes (n = 134). Other reported causes included noise induced (n = 48), sudden deafness (n = 43), explosion or firing practice (n = 40), congenital causes (n = 16), and various other factors (e.g., ear surgery, hereditary factors, tinnitus, infections, concerts, medication, injury, otosclerosis; n = 2–15).

#### General health and lifestyle

Individuals provided information about their overall health, including any limitations in daily activities due to health problems, how their physical and mental health had affected task performance in the four weeks prior to data collection, pain impacting daily activities, emotional well-being, and perceived memory issues. They also reported any chronic conditions diagnosed in their lifetime and in the past 12 months, such as respiratory disorders, cardiovascular diseases, diabetes, musculoskeletal, or psychiatric conditions (see Table 1). Some related conditions were grouped together for brevity in Table 1 (e.g., heart attack, angina pectoris, and cardiac insufficiency as cardiovascular conditions), although they were asked separately to reflect how patients typically recognize these conditions and to enable more detailed information beyond a single general measure. These data are valuable, for example, for assessing the burden of comorbid conditions and for calculating a comorbidity index according to the German National Health Examination Survey^51^. This index quantifies the impact of chronic conditions on the use of health care services, on quality of life, and on other health-related outcomes.

**Table 1.** Prevalence of health problems and hearing difficulties, overall and by gender.

| Health problems | *n* | % in the sample | % of men | % of women |
| --- | --- | --- | --- | --- |
| Hearing Difficulties | 443 | 75.0 | 46.87 | 28.09 |
| Musculoskeletal conditions | 325 | 55.0 | 28.43 | 26.57 |
| Respiratory conditions | 79 | 13.4 | 6.09 | 7.28 |
| Cardiovascular conditions | 77 | 13.0 | 10.49 | 2.54 |
| High blood pressure/cholesterol | 317 | 53.6 | 32.49 | 21.15 |
| Diabetes mellitus | 52 | 8.8 | 5.41 | 3.38 |
| Neurological disease, e.g., Parkinson's disease | 33 | 5.6 | 4.57 | 1.02 |
| Sight disorder/blindness | 83 | 14.0 | 8.46 | 5.58 |
| Risk of falling/dizziness | 75 | 12.7 | 5.08 | 7.61 |
| Chronic gastric-intestinal disease | 65 | 11.0 | 5.41 | 5.58 |
| Cancer | 61 | 10.3 | 5.92 | 4.4 |
| Psychiatric conditions | 71 | 12.0 | 5.58 | 6.43 |
| Other chronic diseases (kidney, bladder, skin, etc.) | 172 | 29.1 | 17.77 | 11.34 |
| Physical handicap (arms/legs) | 108 | 18.3 | 9.31 | 8.97 |

*Note.* Cardiovascular conditions refer to clinically diagnosed disorders affecting the heart or vascular system (e.g., heart attack, cardiac insufficiency or angina pectoris), whereas high blood pressure and cholesterol represent individual cardiovascular risk factors that may occur independently of a formal disease diagnosis. Reporting separately allows for a more refined analysis of the relationship between comorbid health conditions, risk factors, and hearing health.

#### *Short form 12 health survey* (SF-12)^52^

The health status section of the HQ also included the SF-12 health survey, which is a concise version of the 36-item health survey, but comprehensive enough to provide an overview of an individual’s physical and mental health. The SF-12 contains 12 questions, yielding two summary scores. The physical component score (PCS) quantifies limitations in physical functioning, role limitations due to physical health problems, bodily pain, general health perceptions, and social functioning limitations due to physical problems. The mental component score (MCS) indicates vitality, emotional well-being, and social functioning limitations due to emotional problems, as well as interference with daily activities caused by mental health problems. The PCS and MCS raw scores are separately transformed into standardized scores and included in the OHHR as separate scores (*T*-values, with *M = 50, SD= 10)*.

#### Media consumption and device usage

A specific section of the HQ aimed to assess the participant’s media consumption habits and preferences. It included questions about headphone use for TV and radio, detailing the types of hardware and software used (e.g., radio, Bluetooth, infrared) at the time of data collection and how often they were used. Individuals also reported their use of technology for work and personal activities, both with and without hearing aids. In addition, they were asked about their experiences with sound levels, including situations where sounds were perceived as too loud with or without hearing aids. The survey also addressed the demand and perceived sound quality of hearing aids, the frequency of increasing TV volume without them, and individual musical preferences and expectations for sound quality.

#### Technology readiness

The HQ also included a 12-item questionnaire to measure the use of adaptive technology, particularly among older adults^53^. The measurement instrument is based on the theoretical concepts outlined in the technology acceptance model^54^, which postulates how perceived usefulness and ease of use influence technology adoption. Furthermore, it also incorporates concepts from Ajzen’s (1991)^55^ work on personal beliefs about competence and control. Neyer and colleagues (2012)^53^ extended these concepts by developing a comprehensive model of technology readiness that integrates both attitude-oriented and personality-theoretical perspectives. The main construct measured by the 12 items is technology readiness, or the willingness to invest time and effort in learning and using modern technology. Additionally, there are three subscales that can be measured with this questionnaire. Technology competence reflects the perceived ability and confidence in using technology effectively^56^. Technology acceptance quantifies an individual’s relationship with modern technologies, primarily in terms of interest in technical innovations^54^. Technology control reflects the degree to which individuals feel that they have control over their technology use and can overcome challenges^56^.

#### Demographic and socio-economic status

The demographic information was collected in both the clinical interview (age, gender, language status, etc.) and the HQ (household composition and living situation). Participants also reported socio-economic details about themselves, such as their educational degree, profession, occupation, and household income. The Scheuch-Winkler Index (SWI) was used to quantify socio-economic status (SES) by summing up education, occupation, and net income, resulting in a score ranging from 3 to 21^57^. This index is crucial for describing health disparities by investigating how socio-economic factors influence health outcomes. These scores were categorized into three SES groups—low, medium, and high—to represent different socio-economic strata within the German population.

Overall, the HQ collected extensive data that allows for a comprehensive characterization of the impact of hearing loss on an individual’s quality of life and their biopsychosocial characteristics, making this dataset highly valuable for informing precision audiology. By including validated instruments along with newly designed questions, the HQ provided a thorough dataset for analyzing the complex, multivariate interactions between hearing health, general health, technology use, and socio-economic factors. All of this is complemented by further objective measures of hearing and cognition, which are described in the following.

### Clinical interview for anamnesis

To gain a detailed overview about the participants’ hearing health, beyond the survey data recorded with the HQ, an expert interviewer conducted a structured face-to-face interview with the participants. The interview was designed to record further demographic information and questions pertaining to the onset, progression, and specific characteristics of the individual’s hearing loss, along with details on cochlear implants or any other implant, hearing aid use (access and frequency), family history of hearing loss, and history of ear infections.

### Audiological tests

#### Pure Tone Audiometry

Pure tone audiometry is a standard tool used in clinical settings to identify hearing impairments. The resulting audiogram provides a comprehensive assessment of hearing sensitivity, conductive hearing loss, cochlear function, and discomfort levels. The audiometry procedure for OHHR was performed in the laboratory, with each ear assessed independently. Air conduction hearing thresholds were measured using a Siemens Unity II audiometer with Sennheiser HDA200 headphones, while bone conduction thresholds were evaluated with a RadioEar B71 bone conduction transducer. This was done with sinusoids for frequencies ranging from 125 Hz to 8000 Hz in an acoustically shielded audiometry booth. Bone conduction measurements were limited to 500 to 4000 Hz, following standard clinical practice. Below 500 Hz, the vibration threshold is too close to the hearing threshold, making measurements unreliable. Frequencies above 4000 Hz are excluded due to excessive distortion produced by the transducer. A standard clinical threshold method was used with a step size of 5 dB, where a threshold was accepted if the probe tone was not heard twice at lower levels and detected twice at higher levels. To summarize overall hearing sensitivity, the Pure Tone Average (PTA) was calculated as the mean hearing threshold across 500, 1000, 2000, and 4000 Hz. Additionally, measurements of uncomfortable loudness levels (UCL) were performed at 500, 1000, 2000, and 4000 Hz. Tone levels were increased in 5 dB increments and participants were asked to indicate when the sound became uncomfortably loud by pressing a button. The respective presentation level was recorded, and the presentation sequence was continued for the next test frequency at a presumably comfortable level. The complete assessment took approximately 13 minutes.

#### Adaptive Categorical Loudness Scaling

Adaptive Categorical Loudness Scaling (ACALOS) is a method used in clinical audiology to measure an individual’s subjective perception of loudness according to Brand and Hohmann (2002)^42^. It is particularly adept at diagnosing loudness recruitment, which is commonly encountered among individuals with sensorineural hearing loss. This phenomenon is characterized by an excessive increase in perceived loudness as the sound level increases.

The stimulus signals used were one-third-octave bands of noise (duration 2 seconds) presented through Sennheiser HDA 200 headphones for 20 trials in a pseudo-random order. Participants were asked to rate their perceived loudness of each stimulus level using an 11-category response scale, ranging from “not heard” to “too loud”. The categories corresponded to 0, 5, 15, 25, 35, 45 and 50 categorical units, with four unnamed categories situated between them (10, 20, 30, 40 cu). The presentation levels were adaptively adjusted based on the participants’ previous responses, thereby ensuring that the test assessed the whole individual range of loudness perception^42^. Measurements were recorded successively for 1500 and 4000 Hz narrow-band noise stimuli for the left and right ear separately. A loudness function can be fitted to the responses of each of the ACALOS measurements using either the Brand and Kollmeier (2002)^58^ staircase method or the BTUX fitting method^59^, depending on the researcher’s preference^33^.

The ACALOS procedure was designed to provide a reliable and efficient estimation of loudness functions with a small number of trials. It has good test-retest reliability, with intraindividual standard deviations for loudness levels ranging from 4-5 dB, comparable to or slightly better than other procedures that require more trials. The adaptive approach allows for efficient use of trials and better coverage of the auditory dynamic range, particularly benefiting people who are hard of hearing. Additionally, ACALOS eliminates the need for prior measurements of hearing thresholds, simplifying the testing procedure^42^.

While Pure tone audiometry and ACALOS are effective in assessing hearing sensitivity, everyday communication depends on the ability to recognize and understand speech, particularly in noisy environments. Individuals with similar ages and audiogram thresholds can still exhibit substantial differences in speech comprehension in noisy settings. This underscores the importance of including speech-in-noise measures as part of a comprehensive audiological assessment such as the OHHR intends to provide. These additional measures are described below.

#### Digit Triplet Test

The Digit Triplet Test (DTT) employs a closed-set response format that facilitates convenient self-testing via telephone or the internet, primarily for hearing screening purposes. It consists of lists of twenty-seven-digit triplets spoken in background noise used to adaptively determine the speech recognition threshold (SRT). In the adaptive procedure, the signal-to-noise ratio (SNR) for each trial is influenced by the participant’s previous performance. The initial SNR was set at 0 dB, with the noise level remaining constant. The speech presentation level, however, varied: it was decreased following correct identifications and increased if the listener had difficulty understanding any digit^28^. For the current database, the test was administered in the presence of an expert in a controlled laboratory environment. The DTT takes approximately three minutes to complete. The background noise was presented at 65 dB sound pressure level (SPL) for most participants. For those with severe hearing loss, where 65 dB SPL was not sufficiently audible, the presentation level was increased to 80 dB SPL to ensure the test stimuli could be perceived^60^. The purpose of this test is to estimate the SRT, which is defined as the speech level at which a person can hear 50% of the numbers correctly. The DTT offers several advantages, including its brevity, familiarity with the stimuli employed, efficiency, online accessibility, and availability in multiple languages. It has also been shown to be an effective tool for both adults and children with cochlear implants, with of reliability and resilience to learning effects, linguistic abilities, and personal factors such as educational background^61^.

#### Göttingen Sentence Test

The Göttingen Sentence Test (GÖSA) is an open-set speech intelligibility test in which listeners hear 20 meaningful sentences, each presented one at a time in background noise and consisting of three to seven words. The 20-sentence list is randomly selected from a pool of 10 predefined sets of everyday sentences. It provides a more realistic representation of communication situations than tests that have single words or sounds presented in noise. Each sentence is meticulously designed to maintain perceptual equivalence and consistent difficulty levels across different sets, ensuring reliable and comparable results^26,62^.

During the testing session, the sentences were presented with test-specific speech-shaped noise via a free-field loudspeaker in a sound-attenuated room. Participants were instructed to repeat as many parts of the sentence as they could after each sentence presentation. The GÖSA takes about 5 minutes to complete. The noise level during testing is set at 65 dB SPL and increased to 80 dB SPL for participants who were unable to perceive the stimuli adequately at the lower level due to severe hearing loss. The GÖSA also employs an adaptive procedure to determine the SRT with precision in the range of ±1 dB. The noise level is kept constant, and speech levels are adjusted based on the listener’s responses, facilitating accurate assessment of speech perception abilities. In summary, the test offers a comprehensive assessment of speech intelligibility, using ecologically valid materials and carefully calibrated background noise to simulate real-world listening conditions^58^.

### Cognitive measures

Screening tests for dementia and cognitive assessment in general require the participant to hear and comprehend the test items sufficiently well. Similarly, audiological tests, except visual inspection of the ears, demand cognitive abilities such as attention, working memory, and semantic knowledge. These abilities are essential for attending to, comprehending, remembering, executing instructions, and communicating with the examiner. Therefore, an accurate diagnosis of cognitive and audiological conditions is contingent upon a thorough examination of both^63^.

#### Dementia Detection Test

The Dementia Detection test (DemTect) is a neuropsychological screening tool employed to assess cognitive impairment. It is recognized for its high sensitivity and time efficiency, given that it can be administered in 8 to 10 minutes. DemTect comprises five subtests that cover a wide range of cognitive abilities, including immediate and delayed recall of verbal information, working memory, language and number processing, and executive functioning. In the Word List subtest, participants are presented with a 10-item word list over two trials to assess both immediate and delayed recall, thus targeting memory function, which is an essential domain for detecting mild cognitive impairment (MCI) and dementia^64–66^. The Number Conversion task assesses executive function and language processing by requiring participants to switch between different representations of numbers, such as words and numerals. It captures a range of errors, including those related to language impairment, lexical and syntactic processing, and literacy difficulties. Such impairments have been frequently observed in individuals with dementia^67,68^. In the Semantic Fluency Task (’Supermarket’), participants are asked to generate and name items belonging to a specific category, such as supermarket items, within a limited time. It assesses a range of cognitive abilities including attention, working memory, cognitive flexibility, problem solving, semantic memory, language production and processing speed. Studies show that verbal fluency is often impaired in the early stages of dementia^69–72^. The Digit Span task requires participants to repeat sequences of numbers in reverse order and is a measure of working memory. Deficits in this area are considered one of the earliest signs of dementia^25,73,74^. Finally, in the Delayed Recall task, participants are asked to recall the 10-item word list presented at the beginning. This task assesses long-term memory.

The overall score is calculated by summing the scores of each subtest, with each subtest incorporating age-based scoring adjustments for participants under and over 60 years of age to account for age-related differences in cognitive abilities. DemTect has demonstrated robust construct validity and high reliability in both test-retest and inter-rater assessments. Notably, it excels in detecting mild Alzheimer’s disease (AD) with a sensitivity of 100% and mild cognitive impairment with a sensitivity of 80%. In contrast, the widely used Mini-Mental State Examination shows limited efficacy in identifying MCI, with a sensitivity of only 69%^23,25^. This tool enables the identification of potential cognitive decline or impairment of participants in OHHR.

#### Vocabulary size test

The vocabulary size test (in German Wortschatztest-WST^43^) was used to assess the verbal intelligence and language comprehension of the participants included in the OHHR. Factor analytic validation studies have revealed that indicators derived from the vocabulary tests load highly on a general cognition factor (commonly called the *g*-factor), meaning that the scores from WST can be interpreted as effective indicators of *g*^33,43^. Therefore, this assessment has the additional purpose of estimating premorbid intelligence levels in individuals with mild to moderate cognitive impairment and tracking the progression of dementia.

The WST is a 10-minute test consisting of 40-word recognition tasks. In each task, participants must identify the real word presented alongside five similar non-words. The tasks are arranged in a line-by-line format, with increasing difficulty. The raw score is determined by the number of correctly identified words. This test demonstrates high reliability, as evidenced by a split-half reliability coefficient (*r* = .95) and Cronbach’s Alpha (*α* = .94). The score is largely independent of age, exhibiting a very low correlation with age (*r* = .08), but it is positively correlated with educational and vocational qualifications (*r* = .60). It has been normed for a wide age range (20-90 years) and has been standardized using a representative sample (*N* = 572) and Rasch scaling to ensure equivalent measurement of abilities across items.

### Data records

The OHHR is accessible via Zenodo^41^ and is arranged into three principal folders, including **data**, **metadata**, and **tools**. These folders provide all the essential resources needed to access, interpret, and use the dataset easily and effectively. The tools folder contains conversion scripts that read and join the audiogram and loudness scaling JSON tables for potential use in Excel, MATLAB, Python, and SQL, thereby facilitating data management and visualization. The metadata folder includes detailed description files that define all the variables within each .json file, while comprehensive schema files ensure data consistency and enable accurate interpretation across varying platforms. Below is an overview of the principal folders and their structure (see Figure 2), as well as the file names within Table 2.

**Figure 2.**
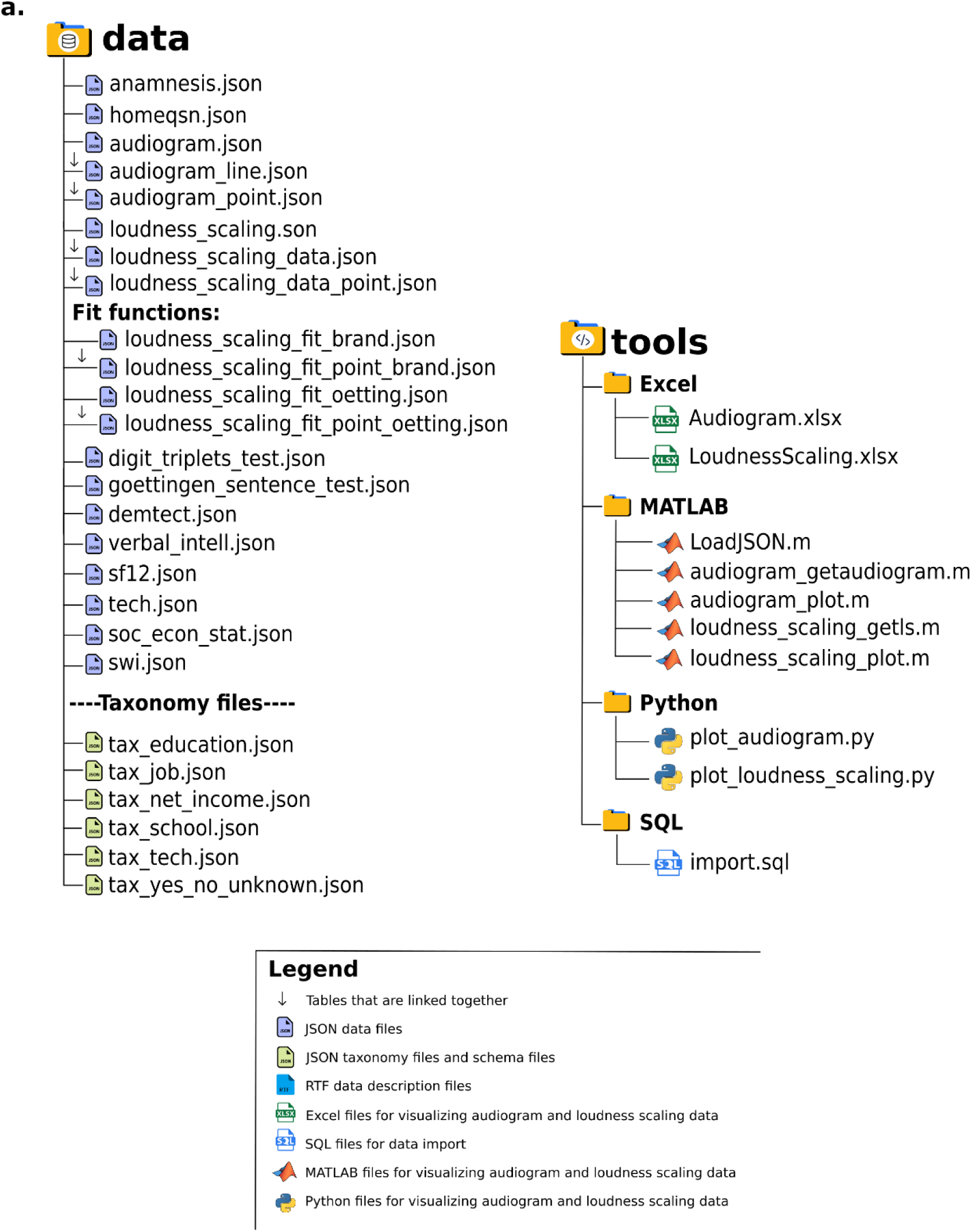
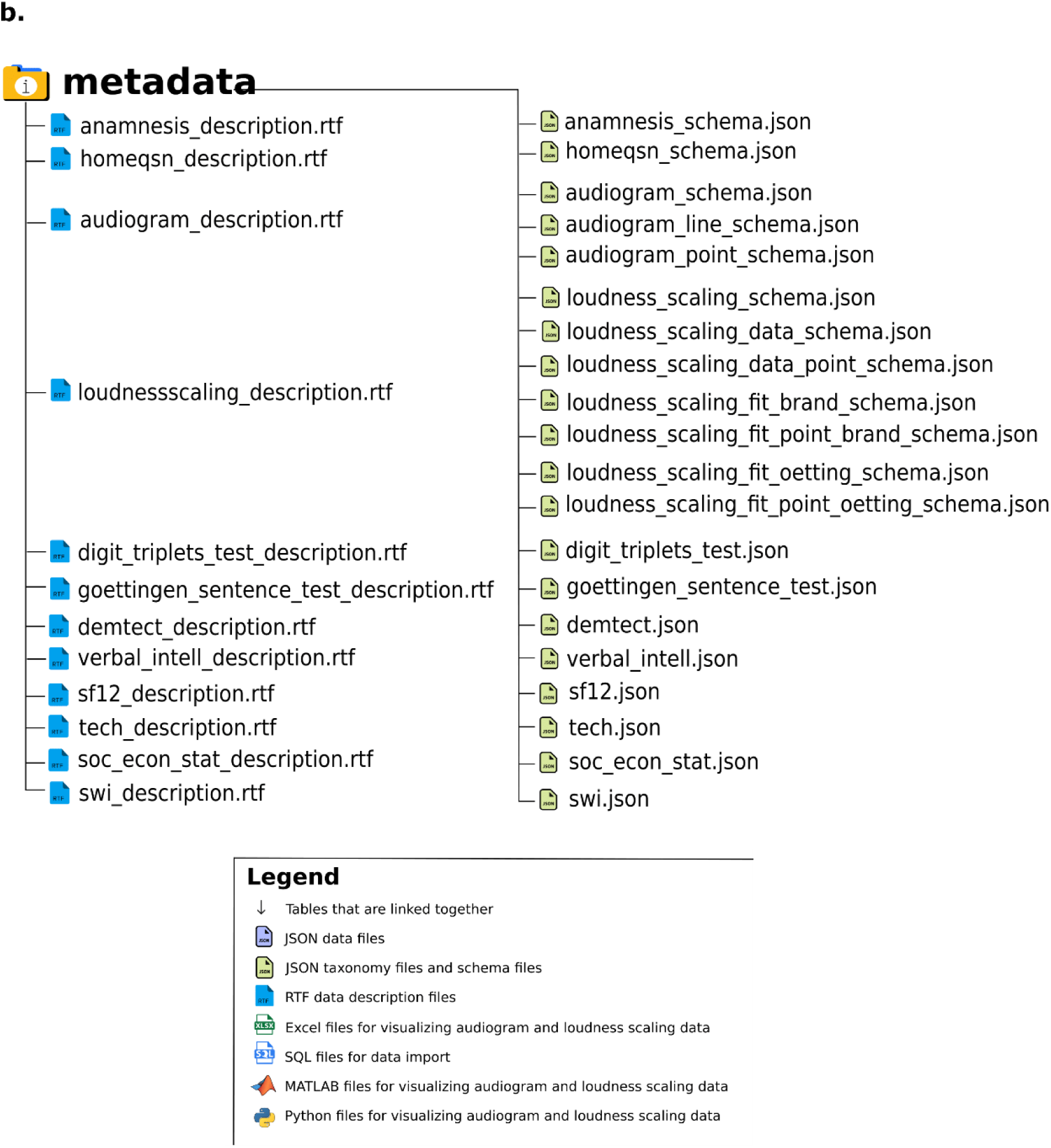
Overview of the data folders and tools of OHHR. *Note*. (a) The data folder contains the core JSON dataset files, including audiogram and loudness scaling tables with linked components (indicated by a downward arrow) that must be joined for use. Taxonomy files define response categories. The tools folder provides resources for data visualization and processing using MATLAB, Python, Excel, and SQL. (b) The metadata folder includes documentation and schema files. RTF files describe each dataset, while JSON schema files define their structure and format.

**Table 2.** Overview of the files providing verbal explanations for levels in categorical data.

| File name | Representation |
| --- | --- |
| tax_education.json | Contains verbal representations of education levels |
| tax_job.json | Contains verbal representations of job levels |
| tax_net_income.json | Contains verbal representations of income levels |
| tax_school.json | Contains verbal representations of school levels |
| tax_tech.json | Contains verbal representations of responses from the technical survey, extracted from the original home questionnaire |
| tax_yes_no_unknown.json | Contains verbal representations for yes, no, and unknown responses |

All the data tables can be found (.json files) inside the data folder and their corresponding schema files in the metadata folder. There are also description files (.rtf format) in the metadata folder that explain the various columns inside each data table. In addition to the primary dataset, the data folder contains six taxonomical .json files that provide verbal representations of different categorical data. The most relevant files are listed and explained in Table 2.

## Technical validation

### Standardized testing protocol

A standardized testing protocol was followed to ensure consistency and reliability in data collection. First, the order and timing of the assessments was standardized. Before any laboratory testing, the HQ was duly completed and returned. The subsequent interviews and assessments were primarily conducted on the same day. However, in certain instances, the assessments were done on alternative days when necessary or to obtain any missing information. To ensure adherence to the established protocol, the dates for each test were recorded. Second, standardized questionnaires and measurements were used wherever possible – see descriptions of the instruments above.

### Medical device certification

The audiometric testing software used was certified as a medical device according to EU regulations and bears the CE mark. Classified as a legacy device under the EU directive, it is suitable for diagnostics in clinical settings and by hearing care professionals. This certification guarantees compliance with stringent safety and performance standards. More details can be found on the <u>Hörzentrum Oldenburg website</u>.

### Calibration procedures

The audiometers were calibrated according to established standards and guidelines (ANSI S3.6-2010; ISO 389-8-2004), ensuring the precision and consistency of our auditory measurements. The speech tests and ACALOS were conducted using the Research and Development version of the Oldenburg Measurement Applications (OMA), designed for scientific research. This software operates on a commercially available PC with a high-quality sound card, supporting multi-channel capability, speech tests in various languages, and customizable noise signals. More details can be found on the <u>Hörzentrum Oldenburg website</u>.

### Data completeness

During the validation process, we thoroughly inspected missing data across both auditory and non-auditory variables. All critical measures and questionnaires were complete, except for SF-12, which had 13 missing entries. The SF-12 was part of the HQ filled out by participants without supervision. Some entries are missing, especially in free response sections if the questions were not relevant to the participant.

### Data quality assessment

We have implemented a comprehensive technical validation process to ensure the highest standards of data quality and reliability. This section details the applied methods and validation steps, emphasizing compliance with prior established knowledge.

First, where applicable, comparisons with normative data were completed to contextualize the study results within established benchmarks or reference points. For instance, the noise level was calibrated to 80 dB SPL during speech-in-noise testing for participants with severe hearing loss who could not adequately perceive the standard 65 dB SPL. Additionally, age-specific scoring criteria were applied in the DemTect assessment to account for age-related changes in cognition. Supplementary Table 1 presents the descriptive statistics for the standardized measurements in the dataset, including the number of items, measurement intention, mean (±SD), and range of values. Normative reference data are primarily available from normal-hearing population samples. The measurements are sensitive enough to differentiate between normal hearing and impairment. However, normative data for specific classes of hearing impairment in the German population were not consistently available in the literature; such data, wherever found, were included in the table^75–77^.

Second, the univariate and bivariate distributions of the audiological measures were analysed according to self-reported normal hearing status or a professional diagnosis of hearing loss. This analysis was conducted to validate that the audiological measures in the OHHR effectively capture group differences. Figure 3 illustrates these distributions, showing clear differences in mean values between groups. Additionally, the figure demonstrates approximately Gaussian distributions within groups and as expected, strong bivariate associations between different measures of hearing ability, supporting the reliability and validity of the audiological data. It is important to note that this figure is intended solely to validate the consistency of hearing-related measures, rather than to assess differences in other audiological or non-audiological variables between groups.

**Figure 3.**
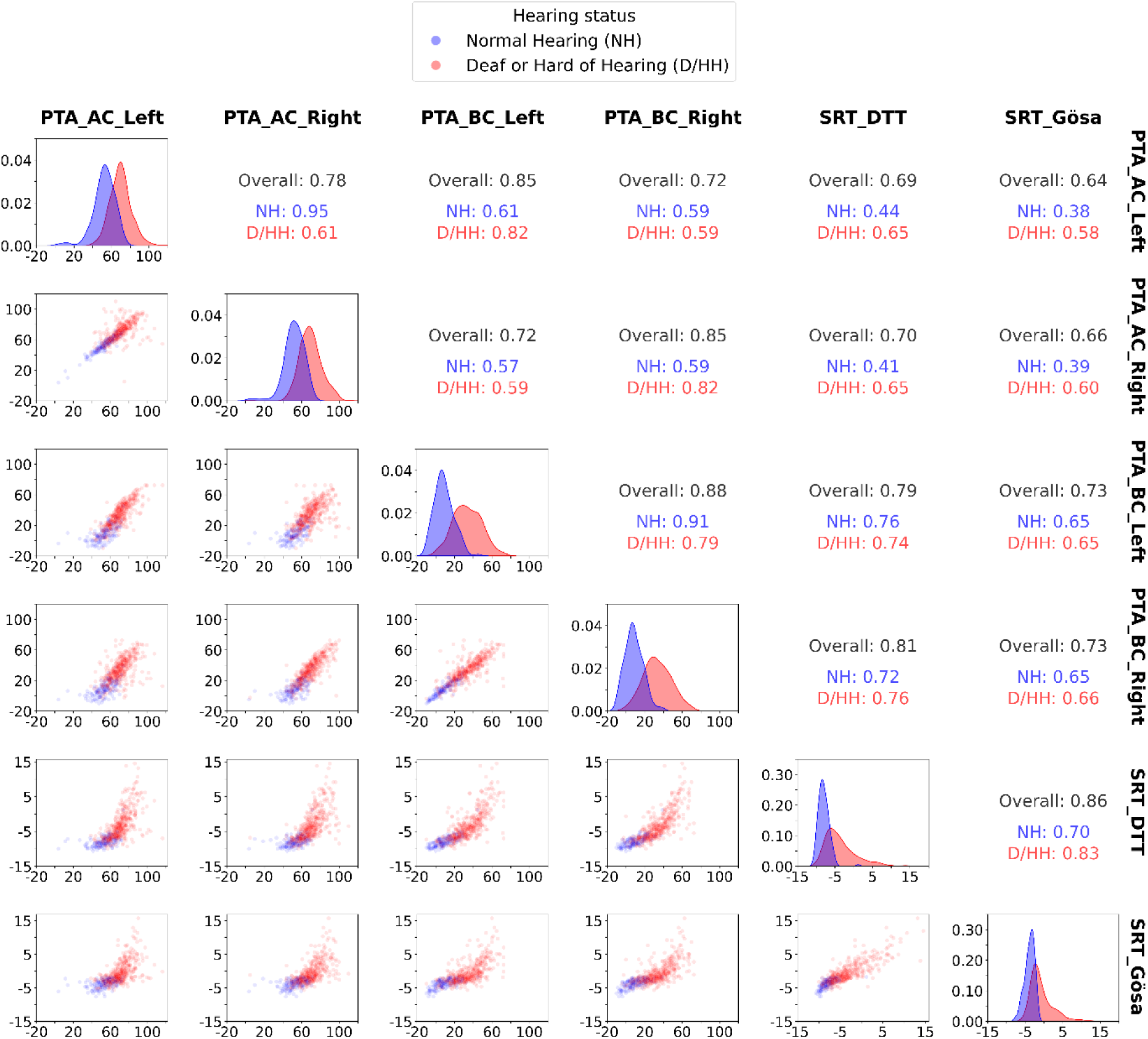
Univariate and bivariate distributions of audiological measures stratified by hearing status. *Note:* PTA (Pure Tone Average): Measures hearing sensitivity across frequencies (0.5, 1, 2, 4 kHz) using air (AC) and bone conduction (BC) thresholds for both left and right ears, expressed in decibels Hearing Level (dB HL). SRT_DTT (Speech Reception Threshold – Digit Triplet Test): Assesses the ability to understand digit sequences in background noise at 50% accuracy, expressed in decibels Signal-to-Noise Ratio (dB SNR). SRT GÖSA (Speech Reception Threshold – Göttingen Sentence Test): Evaluates speech recognition at 50% accuracy with sentences presented in noise, expressed in dB SNR. Off-diagonal plots display the bivariate scatterplots between each pair of measures for normal-hearing (NH; blue) and deaf or hard of hearing (D/HH; red) individuals. Both axes represent the respective measurement values in dB HL for PTA and dB SNR for DTT and GÖSA. Diagonal plots show the distribution of each measure by hearing status, with the x-axis representing measurement values (in dB HL or dB SNR) and the y-axis representing probability density. Correlation coefficients (Pearson’s *r*) for each pair are shown in the upper triangle: Overall, NH, and D/HH., illustrating their relationships within and across groups.

Third, we explored whether the expected low, but well-established association between examiner-assessed and self-reported measures of hearing loss, as reported in the literature, was also present for the variables included in the OHHR. To this end, Figure 4 illustrates the distribution of Pure Tone Average thresholds, as well as Speech Recognition Thresholds from the Göttingen Sentence Test and Digit Triplet Test, across varying levels of self-reported hearing problems. These measures are presented for both quiet and noise conditions. The figure demonstrates a small but consistent positive association between self-reported hearing difficulties and objective measures, including bone and air conduction thresholds, as well as speech-in-noise performance. These findings validate the OHHR hearing-related variables against the established literature.

**Figure 4.**
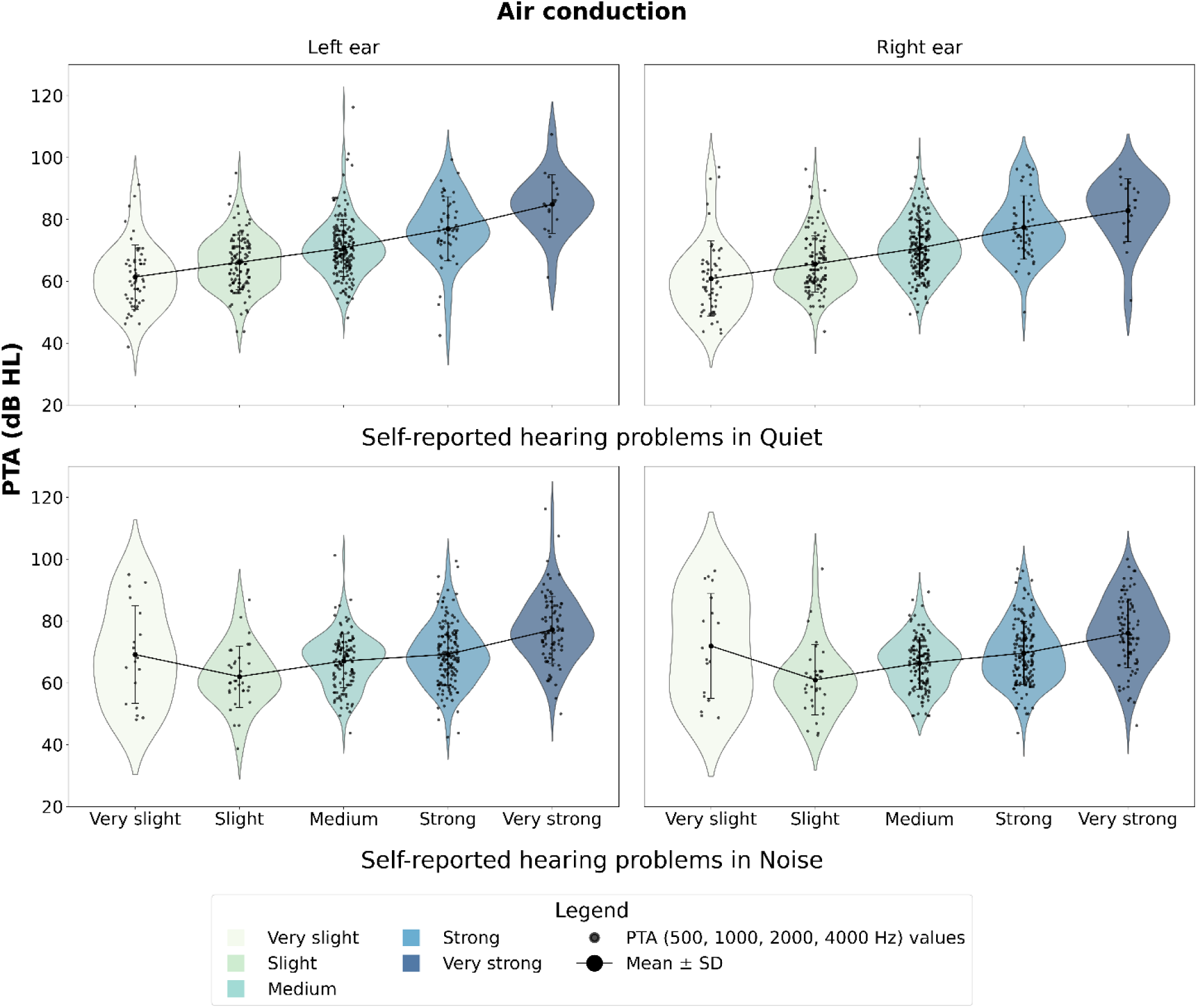
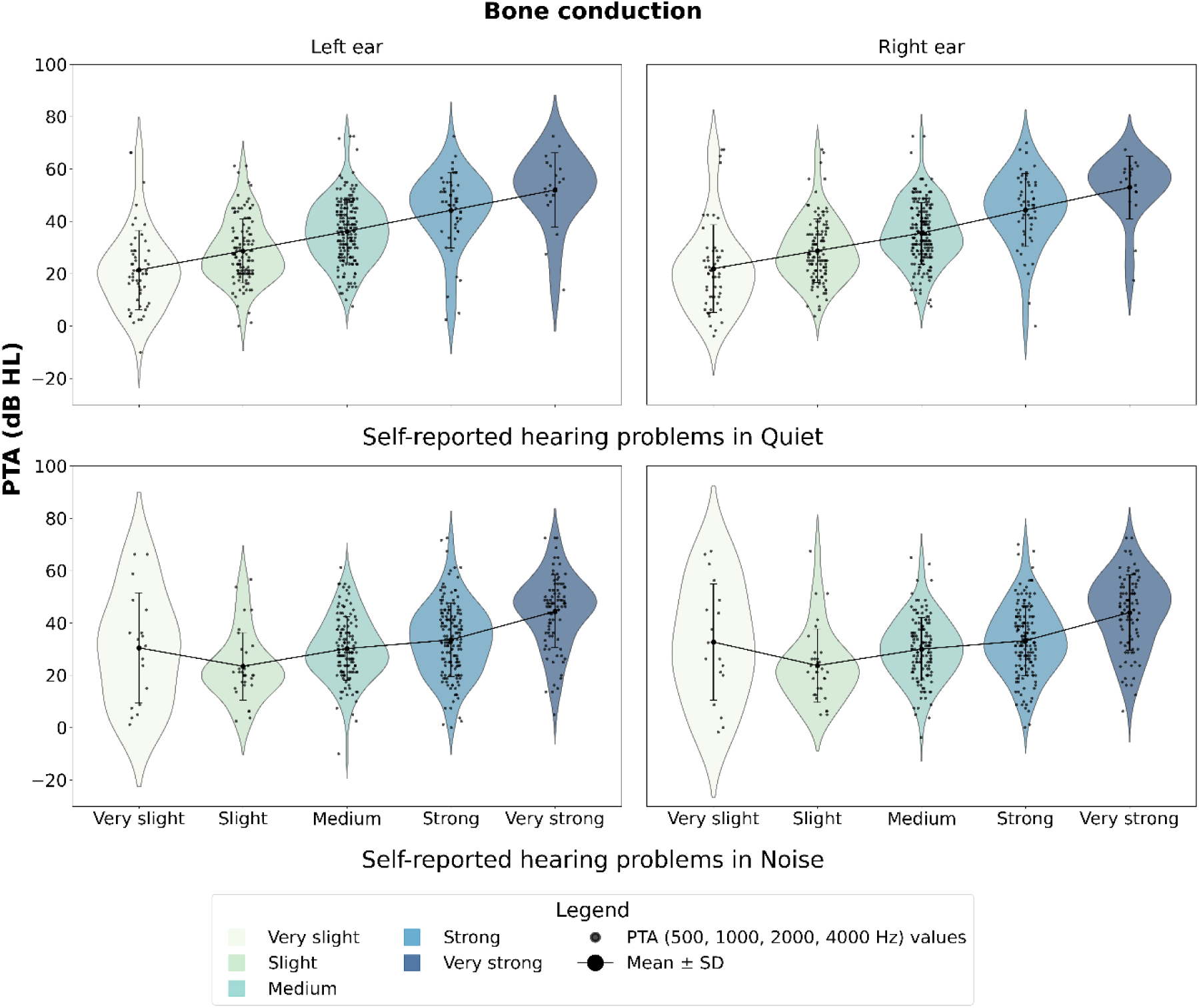
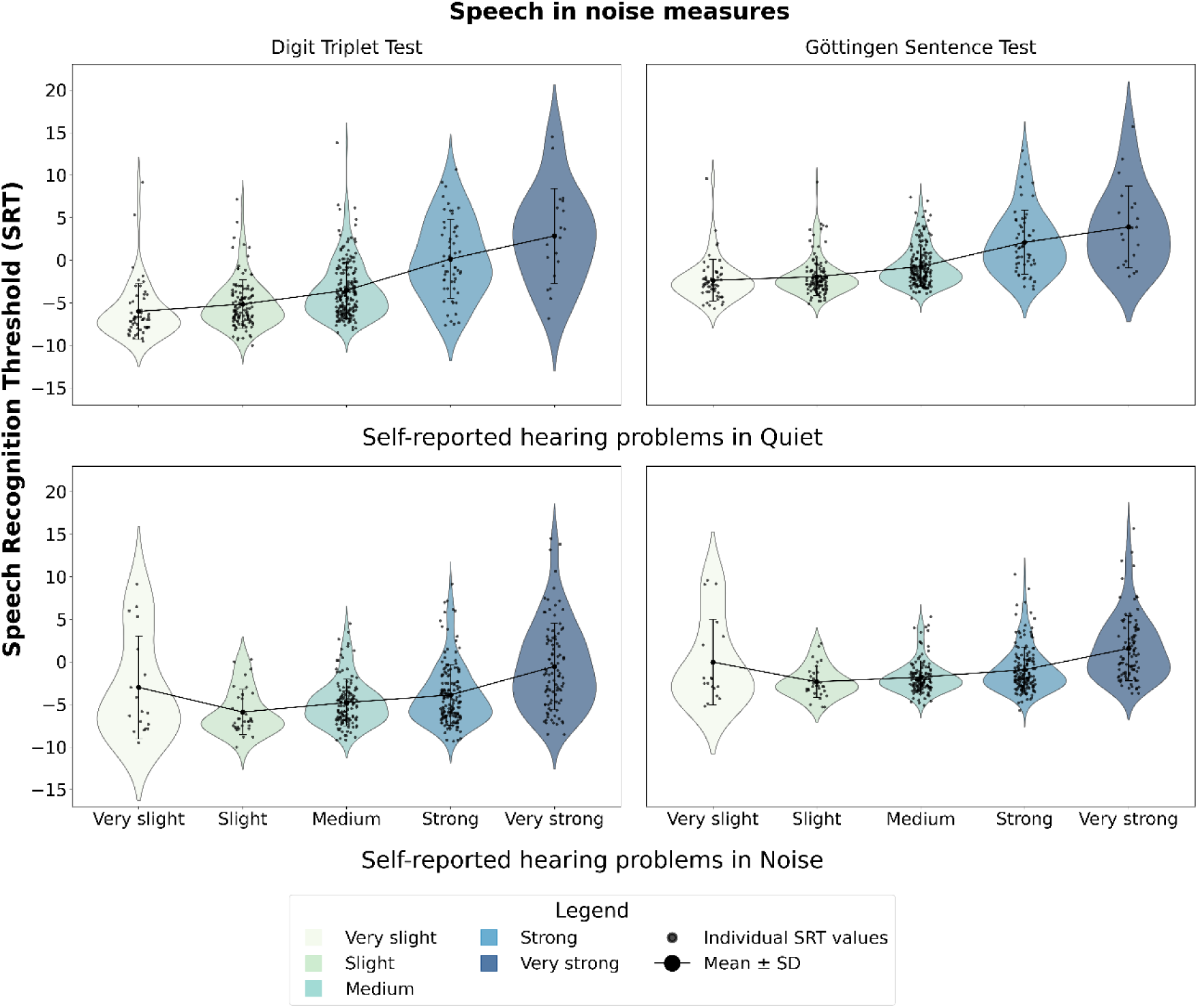
Bivariate distributions of objective auditory measures (PTA Air and Bone conduction, Speech in noise measures) by self-reported hearing severity. *Note.* Comparison of self-reported hearing difficulties and objective auditory measures across three domains: Air Conduction (a), Bone Conduction (b), and Speech-in-Noise Measures (c). The violin plots display the distribution of Pure Tone Average (PTA) thresholds at key frequencies (500, 1000, 2000, and 4000 Hz) for both ears, measured through bone and air conduction, as well as Speech Recognition Thresholds (SRT in dB SNR) from the Digit Triplet Test (DTT) and Göttingen Sentence Test (GÖSA). Self-reported hearing problems are categorized by severity in quiet and noise conditions. Black dots represent individual data points, and solid lines indicate the Mean ± Standard deviation. These plots illustrate how subjective perceptions of hearing difficulties align with objective auditory thresholds and speech-in-noise performance across different tests.

### Representativeness and prevalence

To contextualize the OHHR dataset within the German population, we compared key sample characteristics with those reported in the Gutenberg Health Study (GHS)^2^. The GHS is a large, population-based cohort from Mainz and the Mainz-Bingen district, comprising 5,024 individuals aged 25–86 years (*mean age = 61.2, SD ±13.4 years*; 48.4% women). While the GHS provides population-level estimates, the OHHR intentionally oversampled individuals with hearing loss to capture a broader spectrum of severity for clinical research.

In the OHHR, 76.1% of participants reported being diagnosed with hearing loss, substantially higher than the 34.5% prevalence in the GHS. Applying Röser’s four-frequency classification^78^ to the better ear, the OHHR showed a greater proportion of moderate (22.0%), severe (61.1%), and profound (15.1%) hearing loss, while mild and normal hearing cases were few (1.0% and 0.7%, respectively). In contrast, the GHS reported higher rates of mild loss (22.5%) and lower rates for other more severe categories. Bilateral hearing loss affected 86.6% of OHHR participants, compared to 28.5% in the GHS, and 47% of OHHR participants reported tinnitus, almost double the 25.5% prevalence in the GHS. Bilateral hearing aid provision was also higher (41.8% vs. 7.7%). Sex-specific differences were consistent with the GHS findings, showing hearing loss more common in men (84.7%) than women (65.1%). Hearing loss prevalence in the OHHR increased with age, rising from 44.3% in participants under 55 years to over 80% in those aged 75 and older. This pattern is similar to the age-related trends reported in the GHS, where prevalence ranged from 16.7% in the 55–59 year group to 71.1% in the 75–79 year group.

In the subgroup of participants who reported being diagnosed with hearing loss (n = 442), most belonged to the middle (n = 223) and high (n = 158) socioeconomic status (SES) categories as classified by the Scheuch-Winkler Index, with fewer individuals in the low SES group (n = 61).

Profound and severe hearing loss was more common across all SES levels: profound hearing loss affected 54.1% (low SES), 50.7% (middle SES), and 51.3% (high SES), while severe hearing loss ranged from 29.5% (low SES) to 38.6% (middle SES) and 34.8% (high SES). Unlike the GHS, which reported significantly higher hearing loss prevalence in lower SES groups (p < 0.001), the OHHR cohort showed no clear SES gradient. This likely reflects the clinical nature of the sample and the greater likelihood of individuals from more advantaged backgrounds to participate in research. A similar pattern emerged for education. Among participants with hearing loss, most had middle-level education (n = 279; school students or those with technical or vocational training) or high-level or university education (n = 142), while only 21 individuals had low or no formal vocational qualification. Profound hearing loss was slightly more common in the low education group (54.1%), but both middle (50.5%) and high (51.4%) education groups also showed substantial proportions of profound hearing loss. The small size of the low education group warrants caution in interpreting this difference.

Overall, while our cohort is not representative of the general population in terms of hearing loss prevalence, it is highly representative of the clinical population most affected by hearing impairment. The inclusion of individuals with a broad range of severity, a high prevalence of bilateral hearing loss, and frequent comorbid tinnitus makes this dataset particularly valuable for research on hearing rehabilitation and patient-centered care. The comparison with the GHS highlights these distinctions and situates our findings within the broader context of hearing health in Germany.

## Usage notes

We recommend verifying the completeness of any columns of interest before analysis. If missing data is identified, it is advisable to either impute the missing cases^79^ or exclude them to ensure accuracy.

### Documentation and version control

Comprehensive documentation of data collection methods, transformations, and version control procedures have been prepared. This ensures transparency and accuracy in handling the dataset throughout its lifecycle. Users can access the latest version of the dataset on Zenodo^41^ (see below for access information), where all version histories are recorded.

### Compliance with the FAIR principles

The dataset adheres to the FAIR principles (Findable, Accessible, Interoperable, and Reusable), ensuring usability for a wide range of research. Findability is achieved through its deposition in a public repository such as the “Hearing4all” Zenodo Community, with the present description as a supporting reference. Accessibility is ensured through its open-access status (under CC BY 4.0 license), allowing unrestricted access to all data files, which are stored in machine-readable JSON format and accompanied by comprehensive documentation. Interoperability is supported by storing all data in JSON format including JSON schema files and taxonomies, providing standardized data structures and definitions to facilitate integration with various analytical systems and tools. Reusability is ensured by rich metadata, clear licensing, and custom tools that support data access across popular analysis platforms (Python, MATLAB, Excel, SQL), supporting easy adaptation for future research.

### Temporal consideration and outlook

The data included in the OHHR were collected between 2013 and 2015 and should therefore be interpreted in the context of the period in which they were acquired. Core clinical methods, such as pure-tone audiometry, have remained consistent over time and continue to be widely applied in both clinical practice and hearing research. As a result, these clinical measures do not become outdated as quickly as some research methodologies, which tend to evolve more rapidly. Other components of the dataset, including sentence recognition thresholds in noise and categorical loudness scaling, were collected using methods that were relatively new at the time but have since become more firmly established in hearing research and, to some extent, in clinical audiology. A persistent challenge in expanding the clinical application of such measures are the limited availability of comprehensive reference data from well-characterized hearing-impaired populations, which is precisely the gap the OHHR aims to address.

Although no substantial changes in unaided hearing patterns are currently known, users of the dataset should remain mindful of potential demographic shifts, advancements in hearing aid technology, and evolving healthcare practices that may influence the interpretation and applicability of findings to contemporary populations. Future research could benefit from combining the OHHR with more recent datasets to examine such trends over time, potentially offering valuable insights into changing patterns in hearing health and related outcomes.

## Data Availability

All data collected in this study and described in the preprint is available for open access at Zenodo under the "Hearing4all" Community.

https://zenodo.org/records/14177902

## Code availability

The dataset is available in a machine-readable JSON format, ensuring easy accessibility across different platforms. Custom example scripts are provided to access individual audiogram and loudness scaling files from Python, MATLAB, and Excel, allowing users to analyze and visualize the data effortlessly. Additionally, an SQL script is available for importing the entire dataset into an SQL database for advanced querying. These tools support smooth integration into a wide range of analysis environments.

Excel scripts

- **Excel/Audiogram.xlsx**: An example for using audiograms in Excel. Reads the JSON tables (live data connection), joins the tables as needed, and shows audiogram plots with the possibility to select an observation by its identification number (ID). The relative path of the .json files can be configured on the ‘Setup’ sheet.
- **Excel/LoudnessScaling.xlsx**: An example for using loudness scaling measurements in Excel. Reads the JSON tables (live data connection), joins the tables as needed, and shows loudness function plots with raw data and fits with the possibility to select an observation by ID.

MATLAB scripts

**MATLAB/audiogram_getaudiogram.m:** Returns an audiogram for a user specified observation by joining relevant tables.
**MATLAB/audiogram_plot.m**: Example script for calling ‘audiogram_getaudiogram.m’ and plotting the audiogram.
**MATLAB/loudness_scaling_getls.m**: Returns a loudness scaling measurement for a passed observation by joining relevant tables.
**MATLAB/loudness_scaling_plot.m**: Example script for calling ‘loudness_scaling_getls.m’ and plotting up to four loudness functions with up to two fits and the raw data points. Python scripts
**Python/plot_audiogram.py**: A script for plotting audiograms for an observation by joining relevant tables.
**Python/plot_loudness_scaling.py**: A script for plotting loudness scaling for an observation by joining relevant tables.

SQL script

- **SQL/import.sql**: A script including documentation for importing the database into an SQL Database.

The full dataset, metadata and the above-mentioned scripts are available under CC BY 4.0 license at https://zenodo.org/records/14177902.

## Acknowledgements

We extend our gratitude to all participants who generously contributed their data for the development of OHHR. We also acknowledge Anja Gieseler and Maike Tahden for their efforts in consolidating the specification document detailing the data codes in the previous version of this dataset, which significantly facilitated the description and refinement of the database. This research has been funded by the Deutsche Forschungsgemeinschaft (DFG, German Research Foundation) under Germany’s Excellence Strategy – EXC 2177/1 - Project ID 390895286.

## Author contributions

Conception, design, and preparation of the manuscript: Sumbul Jafri, Daniel Berg, Christiane Thiel, Andrea Hildebrandt

Contribution to study design: Kirsten C. Wagener, Matthias Vormann, Birger Kollmeier

Recruiting participants and data acquisition: Matthias Vormann, Kirsten C. Wagener

Database management: Daniel Berg, Matthias Vormann, Mareike Buhl, Samira Saak

All authors critically reviewed and approved the final version of the manuscript.

## Rights and permissions

**Open Access.** This article is licensed under the Creative Commons Attribution 4.0 International License (CC BY 4.0). This license allows for unrestricted use, sharing, adaptation, distribution, and reproduction in any medium or format, provided that appropriate credit is given to the original author(s) and the source, a link to the license is provided, and any changes made are clearly indicated. All figures included in this article are licensed under the same terms and can be credited with a single line. To view a copy of this license, visit http://creativecommons.org/licenses/by/4.0/

## References

1. World Health Organization. World Report on Hearing. (2021).

2. Döge, J. & others. The Prevalence of Hearing Loss and Provision with Hearing Aids in the Gutenberg Health Study. Deutsches Ärzteblatt International 120, 99 10.3238/arztebl.m2022.0385 (2023).

3. Hackenberg, B. & others. Hearing loss and its burden of disease in a large German cohort—hearing loss in Germany. The Laryngoscope 132, 1843–1849 10.1002/lary.29980 (2022).

4. Arlinger, S. Negative consequences of uncorrected hearing loss—a review. International Journal of Audiology 42, 17–20 10.3109/14992020309074639 (2003).

5. Ray, J., Popli, G. & Fell, G. Association of cognition and age-related hearing impairment in the English longitudinal study of ageing. JAMA Otolaryngology–Head & Neck Surgery 144, 876–882 10.1001/jamaoto.2018.1656 (2018).

6. Wang, H. F. & others. Hearing impairment is associated with cognitive decline, brain atrophy and tau pathology. EBioMedicine 86, 10.1016/j.ebiom.2022.104336 (2022).

7. Pichora-Fuller, M. K. & others. Hearing impairment and cognitive energy: The framework for understanding effortful listening (FUEL). Ear and Hearing 37, 5S–27S 10.1097/AUD.0000000000000312 (2016).

8. Rosemann, S. & Thiel, C. M. The effect of age-related hearing loss and listening effort on resting state connectivity. Scientific Reports 9, 2337 10.1038/s41598-019-38816-z (2019).

9. Rönnberg, J., Holmer, E. & Rudner, M. Cognitive hearing science and ease of language understanding. International Journal of Audiology 58, 247–261 10.1080/14992027.2018.1551631 (2019).

10. Shukla, A. & others. Hearing loss, loneliness, and social isolation: a systematic review. Otolaryngology–Head and Neck Surgery 162, 622–633 (2020).

11. Nordvik, Ø. & others. Generic quality of life in persons with hearing loss: a systematic literature review. BMC Ear, Nose and Throat Disorders 18, 1–13 10.1186/s12901-018-0051-6 (2018).

12. Hornsby, B. W. Y. The effects of hearing aid use on listening effort and mental fatigue associated with sustained speech processing in adults with hearing loss. Ear and Hearing 34, 523–534 10.1097/AUD.0b013e31828003d8 (2013).

13. Dixon, P. R. & others. Health-Related Quality of Life Changes Associated With Hearing Loss. JAMA Otolaryngology–Head & Neck Surgery 146, 630–638 10.1001/jamaoto.2020.0674 (2020).

14. Sethukumar, P. & others. Cataloging Existing Hearing Loss Cohort Data to Guide the Development of Precision Medicine for Sensorineural Hearing Loss: A Systematic Review. International Archives of Otorhinolaryngology https://doi.org/10.5152/iao.2023.22690 (2023) doi:10.5152/iao.2023.22690.

15. Kanatani, Y. & others. National registry of designated intractable diseases in Japan: present status and future prospects. Neurologia Medico-Chirurgica 57, 1–7 10.2176/nmc.st.2016-0135 (2017).

16. Nosrati-Zarenoe, R., Hansson, M. & Hultcrantz, E. Assessment of diagnostic approaches to idiopathic sudden sensorineural hearing loss and their influence on treatment and outcome. Acta Oto-Laryngologica 130, 384–391 10.3109/00016480903161541 (2010).

17. Fink, N. E. & others. Childhood development after cochlear implantation (CDaCI) study: Design and baseline characteristics. Cochlear Implants International 8, 92–116 10.1002/cii.333 (2007).

18. Sudlow, C. & others. UK biobank: an open access resource for identifying the causes of a wide range of complex diseases of middle and old age. PLoS Medicine 12, e1001779 10.1371/journal.pmed.1001779 (2015).

19. Brown, M. 1970 British cohort study. Open Health Data 2, e6 (2014).

20. Fisher, D. E. & others. Sex-specific predictors of hearing-aid use in older persons: The age, gene/environment susceptibility-Reykjavik study. International Journal of Audiology 54, 634–641 10.3109/14992027.2015.1024889 (2015).

21. Ikram, M. A. & others. The Rotterdam Study. Design update and major findings between 2020 and 2024. European Journal of Epidemiology 39, 183–206 10.1007/s10654-023-01094-1 (2024).

22. Xu, P. & others. Hypothalamic substructural integrity is associated with age, sex and cognitive function across lifespan: A comparative analysis of two large population-based studies. https://doi.org/10.1002/alz.084672 (2024) doi:10.1002/alz.084672.

23. Pinto, T. C. & others. Is the Montreal Cognitive Assessment (MoCA) screening superior to the Mini-Mental State Examination (MMSE) in the detection of mild cognitive impairment and Alzheimer’s Disease (AD) in the elderly? International Psychogeriatrics 31, 491–504 10.1017/s1041610218001370 (2019).

24. Tang-Wai, D. F. & others. CCCDTD5 recommendations on early and timely assessment of neurocognitive disorders using cognitive, behavioral, and functional scales. Alzheimer’s Dementia (NY) 6, 10.1002/trc2.12057 (2020).

25. Kalbe, E. & others. DemTect: a new, sensitive cognitive screening test to support the diagnosis of mild cognitive impairment and early dementia. International Journal of Geriatric Psychiatry 19, 136–143 10.1002/gps.1042 (2004).

26. Kollmeier, B. & Wesselkamp, M. Development and evaluation of a German sentence test for objective and subjective speech intelligibility assessment. Journal of the Acoustical Society of America 102, 2412–2421 10.1121/1.419624 (1997).

27. Smits, C., Kapteyn, T. S. & Houtgast, T. Development and validation of an automatic speech-in-noise screening test by telephone. International Journal of Audiology 43, 15–28 10.1080/14992020400050004 (2004).

28. Zokoll, M. A., Wagener, K. C., Brand, T., Buschermöhle, M. & Kollmeier, B. Internationally comparable screening tests for listening in noise in several European languages: The German digit triplet test as an optimization prototype. International Journal of Audiology 51, 697–705 10.3109/14992027.2012.690078 (2012).

29. Van den Borre, E., Denys, S., van Wieringen, A. & Wouters, J. The digit triplet test: a scoping review. International Journal of Audiology 60, 946–963 10.1080/14992027.2021.1902579 (2021).

30. Lehrl, S., Triebig, G. & Fischer, B. Multiple choice vocabulary test MWT as a valid and short test to estimate premorbid intelligence. Acta Neurologica Scandinavica 91, 335–345 10.1111/j.1600-0404.1995.tb07018.x (1995).

31. Overman, M. J., Leeworthy, S. & Welsh, T. J. Estimating premorbid intelligence in people living with dementia: a systematic review. International Psychogeriatrics 33, 1145–1159 10.1017/S1041610221000302 (2021).

32. McDonough, I. M. & others. Discrepancies between fluid and crystallized ability in healthy adults: a behavioral marker of preclinical Alzheimer’s disease. Neurobiology of Aging 46, 68–75 10.1016/j.neurobiolaging.2016.06.011 (2016).

33. Gieseler, A. & others. Auditory and Non-Auditory Contributions for Unaided Speech Recognition in Noise as a Function of Hearing Aid Use. Frontiers in Psychology 8, 219 10.3389/fpsyg.2017.00219 (2017).

34. Tahden, M. A., Gieseler, A., Meis, M., Wagener, K. C. & Colonius, H. What keeps older adults with hearing impairment from adopting hearing aids? Trends in Hearing 22, 2331216518809737 10.1177/2331216518809737 (2018).

35. Buhl, M., Warzybok, A., Schädler, M. R., Majdani, O. & Kollmeier, B. Common Audiological Functional Parameters (CAFPAs) for single patient cases: Deriving statistical models from an expert-labelled data set. International Journal of Audiology 59, 534–547 10.1080/14992027.2020.1728401 (2020).

36. Buhl, M., Warzybok, A., Schädler, M. R. & Kollmeier, B. Sensitivity and Specificity of Automatic Audiological Classification Using Expert-Labelled Audiological Data and Common Audiological Functional Parameters. International Journal of Audiology 60, 16–26 10.1080/14992027.2020.1817581 (2021).

37. Buhl, M. Interpretable clinical decision support system for audiology based on predicted Common Audiological Functional Parameters (CAFPAs). Diagnostics 12, 463 10.3390/diagnostics12020463 (2022).

38. Saak, S. K., Hildebrandt, A., Kollmeier, B. & Buhl, M. Predicting Common Audiological Functional Parameters (CAFPAs) as Interpretable Intermediate Representation in a Clinical Decision-Support System for Audiology. Frontiers in Digital Health 2, 596433 10.3389/fdgth.2020.596433 (2020).

39. Mousavi, H., Buhl, M., Guiraud, E., Drefs, J. & Lücke, J. Inference and learning in a latent variable model for beta distributed interval data. Entropy 23, 10.3390/e23050552 (2021).

40. Saak, S., Huelsmeier, D., Kollmeier, B. & Buhl, M. A flexible data-driven audiological patient stratification method for deriving auditory profiles. Frontiers in Neurology 13, 959582 10.3389/fneur.2022.959582 (2022).

41. Jafri, S. & others. OHHR – The Oldenburg Hearing Health Record [dataset], version 1.2.0.0. https://doi.org/10.5281/zenodo.15727482 (2025) doi:10.5281/zenodo.15727482.

42. Brand, T. & Hohmann, V. An adaptive procedure for categorical loudness scaling. Journal of the Acoustical Society of America 112, 1597–1604 10.1121/1.1502902 (2002).

43. Schmidt, K.-H. & Metzler, P. Wortschatztest. (Beltz, 1992).

44. World Medical Association. World Medical Association Declaration of Helsinki: ethical principles for medical research involving human subjects. JAMA 310, 2191–2194 10.1001/jama.2013.281053 (2013).

45. Pronk, M., Deeg, D. J. & Kramer, S. E. Explaining discrepancies between the digit triplet speech-in-noise test score and self-reported hearing problems in older adults. Journal of Speech, Language, and Hearing Research 61, 986–999 10.1044/2018_jslhr-h-17-0124 (2018).

46. Wang, D. & others. Analysis of influential factors of self-reported hearing loss deviation in young adults. Journal of Public Health 28, 455–461 (2020).

47. Kamil, R. J., Genther, D. J. & Lin, F. R. Factors associated with the accuracy of subjective assessments of hearing impairment. Ear and Hearing 36, 164–167 10.1097/aud.0000000000000075 (2015).

48. Louw, C., Swanepoel, D. W. & Eikelboom, R. H. Self-Reported Hearing Loss and Pure Tone Audiometry for Screening in Primary Health Care Clinics. Journal of Primary Care & Community Health 9, 2150132718803156 10.1177/2150132718803156 (2018).

49. Gibson, W. K., Cronin, H., Kenny, R. A. & Setti, A. Validation of the self-reported hearing questions in the Irish Longitudinal Study on Ageing against the Whispered Voice Test. BMC Research Notes 7, 361 10.1186/1756-0500-7-361 (2014).

50. Cox, R. M. & Alexander, G. C. The International Outcome Inventory for Hearing Aids (IOI-HA): Psychometric Properties of the English Version. International Journal of Audiology 41, 30–35 10.3109/14992020209101309 (2002).

51. Bellach, B. M., Knopf, H. & Thefeld, W. Der Bundes-Gesundheitssurvey 1997/98. Gesundheitswesen Sonderheft 60, S59–S68 (1998).

52. Ware, J. E., Kosinski, M. & Keller, S. D. A 12-Item Short-Form Health Survey: Construction of Scales and Preliminary Tests of Reliability and Validity. Medical Care 34, 220–233 10.1097/00005650-199603000-00003 (1996).

53. Neyer, F. J., Felber, J. & Gebhardt, C. Entwicklung und Validierung einer Kurzskala zur Erfassung von Technikbereitschaft. Diagnostica 58, 87–99 10.1026/0012-1924/a000067 (2012).

54. Davis, F. D. Perceived Usefulness, Perceived Ease of Use, and User Acceptance of Information Technology. MIS Quarterly 13, 319–340 10.2307/249008 (1989).

55. Ajzen, I. The Theory of Planned Behavior. Organizational Behavior and Human Decision Processes (1991).

56. Krampen, G. Fragebogen Zu Kompetenz-Und Kontrollüberzeugungen (FKK). (Hogrefe, Verlag für Psychologie, 1991).

57. Winkler, J. & Stolzenberg, H. Adjustierung des Sozialen-Schicht-Index für die Anwendung im Kinder-und Jugendgesundheitssurvey (KiGGS). (2009).

58. Brand, T. & Kollmeier, B. Efficient adaptive procedures for threshold and concurrent slope estimates for psychophysics and speech intelligibility tests. Journal of the Acoustical Society of America 111, 2801– 2810 10.1121/1.1479152 (2002).

59. Oetting, D., Brand, T. & Ewert, S. D. Optimized loudness-function estimation for categorical loudness scaling data. Hearing Research 316, 16–27 10.1016/j.heares.2014.07.003 (2014).

60. Wagener, K. C. & Brand, T. Sentence intelligibility in noise for listeners with normal hearing and hearing impairment: Influence of measurement procedure and masking parameters. International Journal of Audiology 44, 144–156 10.1080/14992020500057517 (2005).

61. Cullington, H. E. & Aidi, T. Is the Digit Triplet Test an Effective and Acceptable Way to Assess Speech Recognition in Adults Using Cochlear Implants in a Home Environment. Cochlear Implants International 18, 97–105 10.1080/14670100.2016.1273435 (2017).

62. Kuehnel, V., Kollmeier, B. & Wagener, K. Entwicklung und Evaluation eines Satztests für die deutsche Sprache I: Design des Oldenburger Satztests. Zeitschrift Für Audiologie 38, 4–15 (1999).

63. Lemke, U. Hearing Impairment in Dementia – How to Reconcile Two Intertwined Challenges in Diagnostic Screening. Audiology Research 1, 10.4081/audiores.2011.e15 (2011).

64. Petersen, R. C. & others. Mild cognitive impairment: clinical characterization and outcome. Archives of Neurology 56, 303–308 (1999).

65. Petersen, R. C. & others. Current concepts in mild cognitive impairment. Archives of Neurology 58, 1985–1992 (2001).

66. Petersen, R. C. & others. Practice parameter: Early detection of dementia: Mild cognitive impairment (an evidence-based review) [RETIRED]. Neurology 56, 1133–1142 10.1212/wnl.56.9.1133 (2001).

67. Deloche, G. & Seron, X. From Three to 3: A Differential Analysis of Skills in Transcoding Quantities Between Patients with Broca’s and Wernicke’s Aphasia. Brain 105, 719–733 10.1093/brain/105.4.719 (1982).

68. Seron, X. & Deloche, G. From 4 to four: a supplement to ‘From three to 3’. Brain 106, 735–744 10.1093/brain/106.3.735 (1983).

69. Tröster, A. I., Salmon, D. P., McCullough, D. & Butters, N. A comparison of the category fluency deficits associated with Alzheimer’s and Huntington’s disease. Brain and Language 37, 500–513 10.1016/0093-934x(89)90032-1 (1989).

70. Chertkow, H. & Bub, D. Semantic Memory Loss in Dementia of Alzheimer’s Type: What Do Various Measures Measure? Brain 113, 397–417 10.1093/brain/113.2.397 (1990).

71. Monsch, A. U. & others. Comparisons of verbal fluency tasks in the detection of dementia of the Alzheimer type. Archives of Neurology 49, 1253–1258 10.1001/archneur.1992.00530360051017 (1992).

72. Cerhan, J. H. & others. Diagnostic Utility of Letter Fluency, Category Fluency, and Fluency Difference Scores in Alzheimer’s Disease. Clinical Neuropsychologist 16, 35–42 10.1076/clin.16.1.35.8326 (2002).

73. Belleville, S., Peretz, I. & Malenfant, D. Examination of the working memory components in normal aging and in dementia of the Alzheimer type. Neuropsychologia 34, 195–207 10.1016/0028-3932(95)00097-6 (1996).

74. Baddeley, A. D., Baddeley, H. A., Bucks, R. S. & Wilcock, G. K. Attentional control in Alzheimer’s disease. Brain 124, 1492–1508 10.1093/brain/124.8.1492 (2001).

75. von Gablenz, P. & Holube, I. Hearing loss and speech recognition in the elderly. Laryngo-Rhino-Otologie 96, 759–764 10.1055/s-0043-119388 (2017).

76. Thiele, C., Sukowksi, H., Lenarz, T. & Lesinski-Schiedat, A. Göttinger Satztest im Störgeräusch für verschiedene Gruppen von Schwerhörigkeit. Laryngo-Rhino-Otologie 91, 782–788 10.1055/s-0031-1295419 (2012).

77. Drixler, K., Morfeld, M., Glaesmer, H., Brähler, E. & Wirtz, M. Validierung der messung gesundheitsbezogener lebensqualität mittels des short-form-health-survey-12 (SF-12 Version 2.0) in einer deutschen normstichprobe. Zeitschrift für Psychosomatische Medizin und Psychotherapie 66, 272–286 10.13109/zptm.2020.66.3.272 (2020).

78. Boenninghaus, H. G. & Röser, D. New tables for the determination of percentile loss of speech hearing. Zeitschrift für Laryngologie, Rhinologie, Otologie und ihre Grenzgebiete 52, 153–161 (1973).

79. Jäger, S., Allhorn, A. & Bießmann, F. A benchmark for data imputation methods. Frontiers in Big Data 4, 693674 10.3389/fdata.2021.693674 (2021).

