## Supplementary Table 1 for "The Oldenburg Hearing Health Record (OHHR)"

**Supplementary Table 1. Descriptive statistics of standardized measures in the OHHR and reference data from other studies**

| Assessment tool | Number of items | Measurement intention | OHHR Sample (Mean ± SD; Range) | Published Norms (German population) |
|---|---|---|---|---|
| Digit Triplet Test<br>(DTT, Zokoll et al., 2012)[23] | 27-digit triplets | Speech Recognition Threshold in noise | -4.6 dB ± 4.2;<br>-10.7 to 14.5 (dB) | Normal Hearing[23]:<br>-9.3 ± 0.4 dB (headphones)<br>-6.5 ± 0.4 dB (telephone) |
| Göttingen Sentence Test<br>(GÖSA, Kollmeier & Wesselkamp, 1997)[21] | 20 sentences | Speech Recognition Threshold in noise | -1.5 dB ± 3.1;<br>-7.6 to 15.7 (dB) | (von Gablenz & Holube, 2017)[67]<br>Young Normal Hearing: -4.8 ± 0.9 (dB);<br>(Range: -6.6 to -3 dB)<br>Hörtech Recommended[67]: -6.2 ± 2.0 (dB);<br>(-8.2 to -4.2 dB)<br>(Thiele et al., 2012)[68]<br>Mild Hearing Loss: 0 ± 1-2 (dB);<br>Moderate Hearing Loss: 5 ± 6 (dB);<br>Severe Hearing Loss: >> 20 (dB) |
| Dementia Detection Test (DemTect, Kalbe et al., 2004)[20] | 5 subtests | Cognitive impairment screening | 15.8 ± 2.3;<br>7 to 18 | Normal Hearing[20]:<br>Normal cognition: Scores ≥13;<br>Mild cognitive impairment: 9–12;<br>Suspected dementia: ≤8 |

| | | | | |
|---|---|---|---|---|
| Vocabulary Size Test (WST, Schmidt & Metzler, 1992)[35] | 40 tasks | Indicator for crystallized intelligence | 31.5 ± 4.9;<br>8 to 41 | Normal Hearing[35]:<br>Healthy young adults: *Mean* 31-35 (depending on education) |
| Short Form Health Survey (SF-12, Ware et al., 1996)[44] | | | | Normal Hearing (Drixler et al., 2020)[69]: |
| Physical Component Score | 6 items | Physical health score (Standardized) | 47.2* ± 9.2;<br>18.7 to 62.9* | 50±10; |
| Mental Component Score | 6 items | Mental health score (Standardized) | 52.1* ± 8.9;<br>21.8 to 70.1* | 50±10 |

Note: The SF-12 values are based on 568 participants due to 13 missing entries. The Physical Component Summary (PCS) and Mental Component Summary (MCS) scores were calculated separately from their respective 6 items each, out of the total 12 items. The mean, range, and standard deviation values for the other measures are based on the entire sample of 581 individuals.
